# Clustering of pulmonary tuberculosis in Ethiopia: repeated population-based symptom screening

**DOI:** 10.1101/2024.08.31.24312883

**Authors:** Abiot Bezabeh Banti, Daniel Gemechu Datiko, Brita Askeland Winje, Sven Gudmund Hinderaker, Einar Heldal, Mesay Hailu Dangisso

## Abstract

**Objective:** A “Cluster” is an area with a higher occurrence of tuberculosis than would be expected in an average random distribution of that area. Tuberculosis clustering is commonly reported in Ethiopia, but most studies rely on registered data, which may miss patients who do not visit health facilities or those who attend but are not identified as having tuberculosis. This makes the detection of actual clusters challenging. This study analysed the clustering of pulmonary tuberculosis and associated risk factors using symptom-based population screening in Dale, Ethiopia.

**Desgign:** A prospective population-based cohort study.

**Setting:** All households in 383 enumeration areas were visited three times over 1 year period, at four-month intervals.

**Participants:** Individuals with pulmonary tuberculosis aged ≥15 years with demographic, socioeconomic, clinical, and geographic data residing in 383 enumeration areas (i.e., the lowest unit/village in the kebele, each with approximately 600 residents).

**Outcome measures:** Pulmonary tuberculosis (i.e., bacteriologically confirmed by sputum microscopy, GeneXpert or cluture plus clinically diagnosed pulmonary tuberculosis) and pulmonary tuberculosis clustering.

**Results:** We identified pulmonary tuberculosis clustering in 45 out of the 383 enumeration areas. During the first round of screening, 39 enumeration areas showed pulmonary tuberculosis clustering, compared to only three enumeration areas in the second and third rounds. Our multilevel analysis found that enumeration areas with clusters were located farther from the health centres than other enumeration areas. No other determinants examined were associated with clustering.

**Conclusions:** The distribution of pulmonary tuberculosis was clustered in enumeration areas distant from the health centres. Routine systematic community screening using existing health infrastructure with Health extension workers may be costly but through targeted screening they can identify and refer persons with TB symptoms more quickly for diagnosis and treatment, thereby decreasing the duration of disease transmission and contributing to the reduction of TB burden.

**Stregths and limiations of this study:**

- The study applied a three-round total household symptom screening strategy to identify undiagnosed as well as diagnosed tuberculosis cases, and hence identify real clusters.
- Health extension workers actively involved the entire population in screening, benefitting from their trust and familiarity with the community.
- The study also sought for risk factors for clustering that may require attention from public health practices at the lowest community level.
- Smear microscopy will due to relatively low sensitivity always miss some cases of tuberculosis.

## Background

Although tuberculosis (TB) is preventable and curable, globally an estimated 1.3 million people died from TB in 2022. Modelling shows that 3.1 million TB cases were not detected or reported globally, which sustains disease transmission and suffering in the community (1, 2). According to the World Health Organization, Ethiopia has a high-burden of TB, with estimated incidence rates of 126 per 100,000 population. The TB burden is particularly acute in certain areas, creating a “TB belt” in the southern part of the country. Higher notification rates in the study area than the national average disproportionately affect these communities.

Efforts to improve services to the most vulnerable and disadvantaged populations using existing health systems are among strategic priorities (3, 4). Clustering of TB in an area means there are more cases than expected from random distribution; this represents an increased risk of transmission (5), but can also provide valuable information for the national TB programme. Clusters can be measured using surveillance data, genotypic data, and statistical models (6). Many studies, including some Ethiopian, have reported TB clustering using surveillance data (6–9). These studies relied on notified data from passive case-finding, which may underestimate the actual TB rate because of undetected cases (10, 11). Population-based studies may help better substantiate the existence of clusters, but they are relatively few (3, 6).

A suggested prevention and care strategy is to target the population in clusters (12). Repeated population screening may help identify these clusters, facilitating the design of focused, less expensive and easily accessible interventions. In our study of pulmonary tuberculosis (PTB) prevalence and incidence, we conducted three rounds of household screening in the Dale district in Ethiopia. We discovered that two-thirds of symptomatic PTB patients were undetected at the time of the household visit (13). This study is based on active case-finding data, complementing information from TB registries, resulting in a more comprehensive dataset on PTB. Our objectives in the Dale district of Ethiopia were to: 1) identify symptomatic PTB clustering through population screening, and 2) explore the association between PTB clustering and demographic, socioeconomic, clinical and environmental risk factors.

## Methods

### Study design and setting

This study is based on data from a cohort study with repeated household screening between October 2016 to September 2017, combined with a follow-up survey to collect geographical data in August and September 2018. The target population included everyone residing in the Dale district, which is located in the Sidama region of southern Ethiopia.

According to the 2017 census, the estimated adult population aged 15 years and older in Dale was 136,181. It’s worth noting that the majority of the population in this region resides in rural areas. Dale has 36 kebeles, which are the lowest administrative units. A kebele is divided into 383 smaller villages called enumeration areas (EAs). EA has 600 persons on average. TB health care services are delivered based on the End TB strategy across 10 health centres, 2 clinics, and 36 health posts. However, neither GeneXpert nor X-ray services are available within Dale. Access to these services requires a referral to the hospital in neighbouring Yirgalem town. In Ethiopia, public local community health providers, Health Extension Workers (HEWs), offer cost-effective health services, which include patient referrals and regular household visits. Approximately 33% of rural residents turn to HEWs as their initial source of care when they become ill (14). We followed the Strengthening the Reporting of Observational Studies in Epidemiology cohort reporting guidelines (Supplemenatl file 1) (15).

### Study population

In the three rounds of household visits between 2016 and 2017, the initial visits occurred from October 2016 to January 2017, followed by visits from February to May 2017, and then again from June to September 2017. The visits occurred every 4-month intervals and lasted 1-2 months each. During these visits, trained HEWs went door-to-door, asking all family members if they had any symptoms compatible with TB, they also asked about symptoms concerning household members who were not at home during the screening and revisited households that were missed. We used the term ’’presumptive TB’’ for individuals having a persistent cough for 14 days or more with or without haemoptysis, weight loss, fever, night sweats, chest pain or difficulty breathing. During these visits, demographic, socioeconomic, clinical, and environmental data to assess associated risk factors were collected (13). Those with symptoms were closely followed and asked to come to the health post to provide sputum. Sputum samples were collected and transported to the health facility for smear microscopy. GeneXpert testing was performed on smear-positive samples to validate the results. In total, the screening identified 442 PTB cases, with 90 clinically diagnosed and 352 bacteriologically confirmed cases. In 2018, we conducted a follow-up survey in the same population collecting geographic information on the identified PTB patients. This allowed us to link the previously collected data on risk factors to the geographic data. The addresses of 17 PTB patients from two kebeles (Hida Kaliti and Kaliti Simita) were not found due to industrial zone construction. Further details on household screening and follow-up can be found elsewhere (13,16).

### Operational definitions

Bacteriologically confirmed PTB was defined as any TB diagnosed based on sputum smear-microscopy, culture or GeneXpert laboratory results. Clinically diagnosed PTB was defined by a clinical decision to start TB treatment in those with a persistent cough, usually supported by radiological findings and evidence from other tests according to the national TB and leprosy guidelines (16). An individual diagnosed with bacteriologically confirmed or clinically diagnosed PTB was considered a PTB case. The most likely and secondary clusters were reported by scan statistics (17). Assuming constant risk, the “most likely PTB cluster” within the EAs has the highest probability of having the most cases (18,19). The “secondary PTB cluster” does not overlap with the most likely cluster but has an increased cluster probability (20). Depending on the risk in the cluster secondary clusters are further divided into secondary cluster 1 and 2. Clustering is a combination of the most likely and secondary clusters. To estimate distance we calculated the straight line from the health centres to the patient’s household.

### Patient and public involvement

No patients were involved in setting the research question or the outcomes, interpretation, writing or dissemination of the results.

### Data collection

The geographical information was collected using a handheld Global Positioning System (GPS). We used GPS to measure the location and altitude, computed population density from census data of the EA, and linked it to the geographic data (GPS coordinates) corresponding enumeration areas. We also used GPS points in the enumeration areas to calculate the straight-line distance between each household and the nearby health center. Locations of patients were collected by trained personnel. HEWs who were familiar with the villages provided support to the data collectors. We used principal component analysis to calculate a wealth index (Supplemental file 2). Socioeconomic covariates obtained directly from the study population were assigned wealth-index scores, which were derived from the original variables measuring household socioeconomic status. We used the wealth index to indicate household income inequality. We determined a Body Mass Index (BMI) cut-off point using national TB guidelines (<16 kg/m^2^ refers to severe acute malnutrition; 16-17 kg/m^2^ refers to moderate acute malnutrition, 17-18.5 kg/m^2^ refers to mild malnutrition, and ≥18.5 kg/m^2^ refers to normal) while the Guinea-Bissau study for the Mid-upper Arm Circumference (MUAC) <22cm refers to malnutrition; MUAC ≥ 22 refers to normal (21,22).

We calculated the median values of the dataset. We used the median value when we divided our dataset into two groups for distance, population, population density and altitude. EA served as the spatial unit of analysis. We used Scan statistics to identify clusters. The cluster was the outcome of interest. We classified EAs with PTB clusters as ’’yes’’ while those without as ’’no’’ to facilitate further analysis using the multilevel technique.

### Statistical analysis

Spatial statistics are commonly used to detect risk factors, hotspots (areas with increased risk transmission or burden), and transmission patterns (23). We performed purely spatial analysis using poisson model to identify the most likely and secondary PTB clusters, using Kulldroff’s scan statistics, with the freely available software (SaTScan 10.1) (17). We analysed data after exploration to avoid systematic errors that could occur during data collection and entry that impact our study’s results. For spatial analysis, we used corrected shape file data linked with EAs. We used the population size for each EA projected from the 2007 census and calculated the expected incidence rate of PTB for Ethiopia. We used ArcGIS 10.4 (ESRI 2016) to calculate centroids for the polygons and construct output maps and maps created using the UTM Zone 37 North projected coordinate system. We used case, population, and coordinate as input for the purely spatial analysis. This analysis distributes the number of events in an area according to the at-risk population. Spatial clustering analysis provides the data with a multilevel structure in which individuals are nested in cluster locations, which allows for an exact estimate of the effect. A between-class coefficient of zero means no clustering effects within the data, that is, all people within the same group have identical responses to the outcome variable (24,25). When dealing with multilevel analyses, multiple groups were involved (24). The analysis included 425 individuals with PTB in two-level regression (patient and EA levels). We evaluated random effects at the EA level by using independent covariance and examined independent factors.

We used three models: In Model 1, we evaluated a null model with no independent covariates. In Model 2, we evaluated individual-level covariates. In Model 3, we added EAs- level covariates to Model 2 and allowed observation days to vary across both levels. We used Jamovi (Version 2.3) software for multilevel analysis (26).

### Ethics clearance

Ethics approval was obtained from the Armauer Hansen Reaserch Institute-Alert Ethics Review Committee, Ethiopia (PO12/15), the National Research Ethics Committee, Ethiopia (no 104/2016), and the Regional Committees for Medical and Health Research Ethics in Norway (2015/1006). Written consent was obtained from participants and parent(s)/Guardian(s) for participants aged 15-17 years.

## Results

### Patient characteristics and distribution

From October 2016 to September 2017, 442 PTB patients were identified. Among them, 425 patients were included in this report, while 17 (3.8%) PTB patients were excluded due to missing residential addresses (see Figure 1). The median age of the patients was 28 years, with an age range of 15 to 90 years. The patients’ mean number of observation days was 34 days with a standard deviation of 2.6 days. Of the patients, 52% were male, 72% were ever married, and only 15% had BCG scars. Of 383 EAs in the district, 197 reported at least one PTB patient, and the geographic distribution of PTB patients exhibited variations (Figure 2).

**Figure 1.**
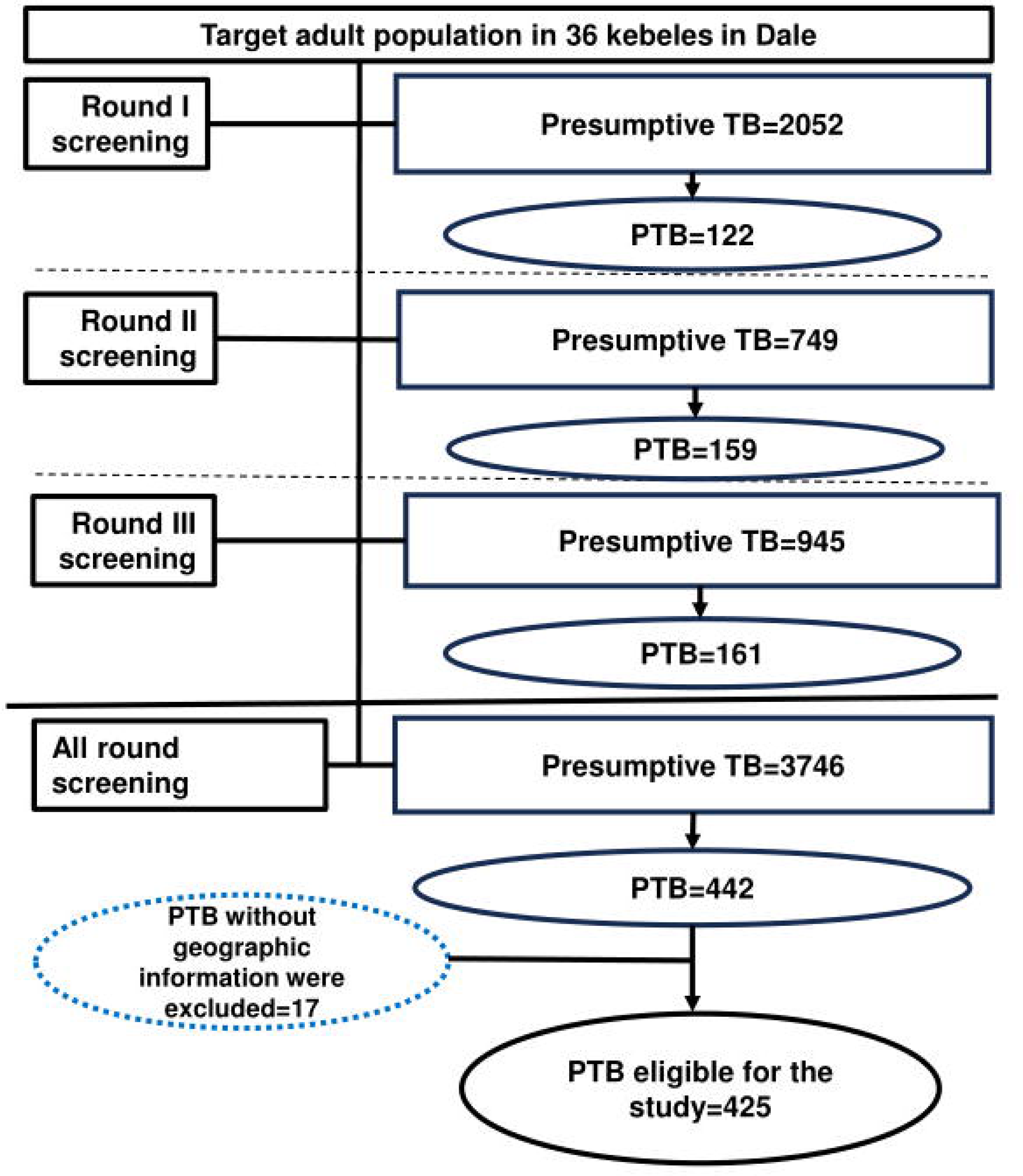
Study flow chart for a household investigation for tuberculosis in Dale district Ethiopia, 2018.

**Figure 2.**
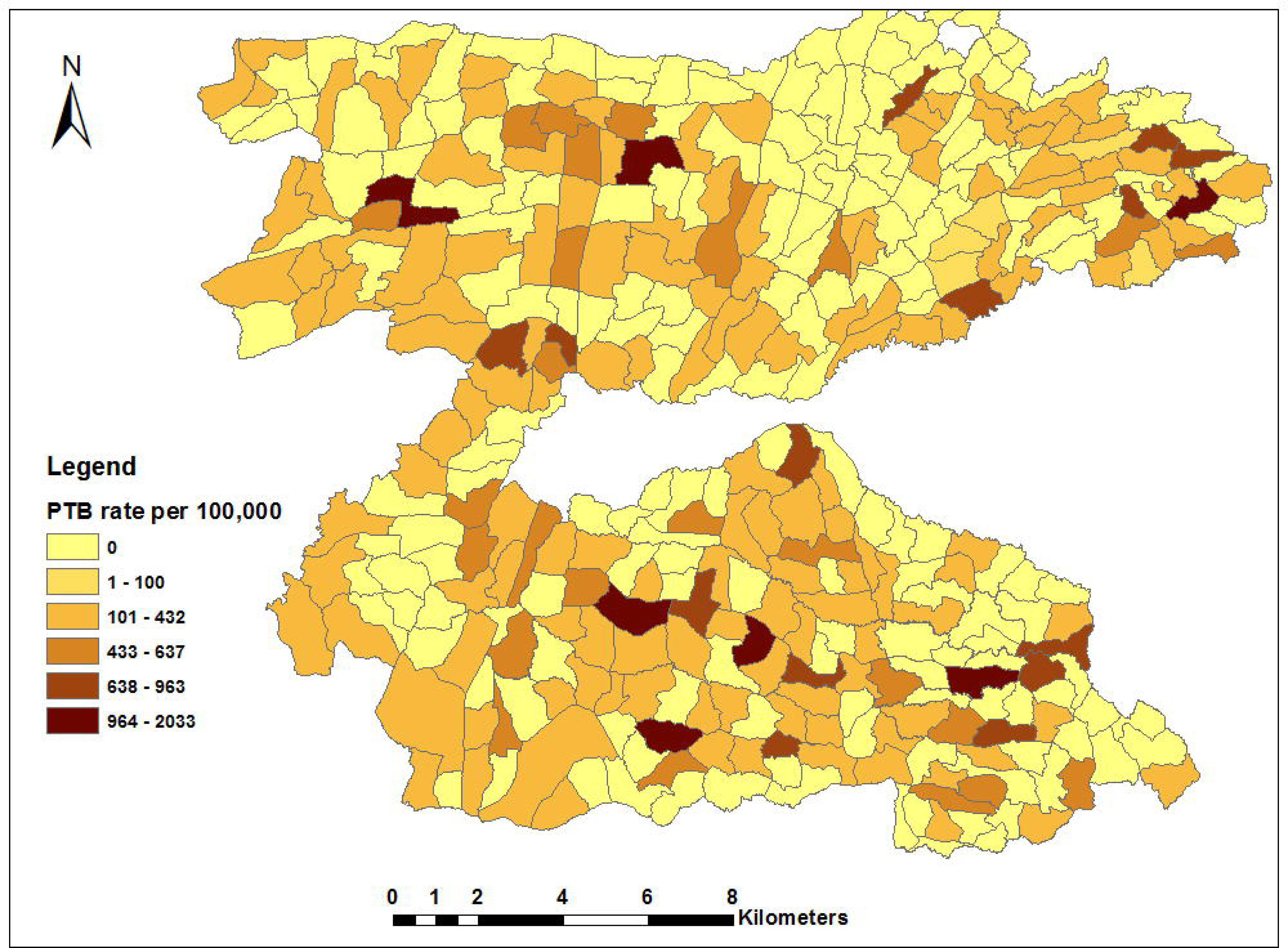
Rate per 100,000 population of pulmonary tuberculosis by Enumeration area in the three rounds of household visits from October 2016 to September 2017, Dale Ethiopia.

Table 1 shows the characteristics of patients inside and outside clusters. Most variables such as age, sex, marital status, BMI, MUAC, Wealth index, and BCG scar, were comparable. Differences appear in schooling (74% vs 59%), distance from health facilities (84% vs 43% in more than 3km), and population density (73% vs 53% in less dense areas <665/square km).

**Table 1.** Characteristics of symptomatic pulmonary tuberculosis patients inside and outside clusters, Dale, Ethiopia 2018.

| Covariate |  | Cluster (n, 425) |  |
| --- | --- | --- | --- |
|  |  | Yes, n (%) | No, n (%) |
| All patients |  | 81 | 344 |
| Sex |  |  |  |
|  | Men | 39 (48) | 180 (52) |
|  | Women | 42 (52) | 164 (48) |
| Age in years |  |  |  |
|  | 15-34 | 60 (74) | 224 (65) |
|  | ≥35 | 21 (26) | 125 (35) |
| Marital status |  |  |  |
|  | Single | 25 (31) | 93 (27) |
|  | Ever married | 56 (69) | 251 (73) |
| Schooling |  |  |  |
|  | Yes | 60 (74) | 204 (59) |
|  | No | 21 (26) | 140 (41) |
| BMI kg/m<sup>2</sup> |  |  |  |
|  | <16 | 22 (27) | 99 (29) |
|  | 16-17 | 12 (15) | 79 (23) |
|  | 17-18.5 | 18 (35) | 80 (23) |
|  | ≥18.5 | 19 (23) | 86 (25) |
| MUAC |  |  |  |
|  | <22 cm | 65 (80) | 267 (78) |
|  | ≥22 cm | 16 (20) | 77 (22) |
| Wealth index |  |  |  |
|  | Poor | 35 (43) | 145 (42) |
|  | Middle | 19 (24) | 106 (31) |
|  | Rich | 27 (33) | 93 (27) |
| BCG scar |  |  |  |
|  | Yes | 13 (16) | 51 (15) |
|  | No | 68 (84) | 293 (85) |
| *Distance from the health centre* |  |  |  |
|  | <3 km | 13 (16) | 197 (57) |
|  | ≥3 km | 68 (84) | 147 (43) |
| *Population density per square km |  |  |  |
|  | <665 | 59 (73) | 183 (53) |
|  | ≥665 | 22 (27) | 161 (47) |
| *Population |  |  |  |
|  | <644 | 57(70) | 196(57) |
|  | ≥644 | 24(30) | 148(43) |
| *Altitude (m) |  |  |  |
|  | <1793 | 57 (70) | 178 (52) |
|  | ≥1793 | 24 (30) | 166 (48) |

BMI, Body Mass Index; MUAC, Mid Upper Arm Circumference; BCG, Bacillus and Calmette
\*These variables used cut-off for groups using the mean values of dataset.

### Clustering of symptomatic PTB

In purely spatial analysis, we found that 45 out of the 383 EAs visisted in each round had PTB clustering during the study period, as shown in Table 2. One EA exhibits the most likely clustering (RR 11.9, p<0.0001), while 41 EAs had secondary cluster 1 (RR 2.09, p<0.001), and 3 EAs had secondary cluster 2 (RR 3.9, p<0.044). PTB clusters were common in the southeast, northwest, and southcentral parts of the district, as shown in Figure 3a-c. These figures show PTB clustering during (a) the entire study period, (b) the first round of household screening, and (c) the second and third rounds of household screening. In the first round, 39 EAs (10%) had PTB clusters, but only three PTB clusters (1%) were detected in two kebeles during the second and third visits between February and September 2017.

**Figure 3a-c.**
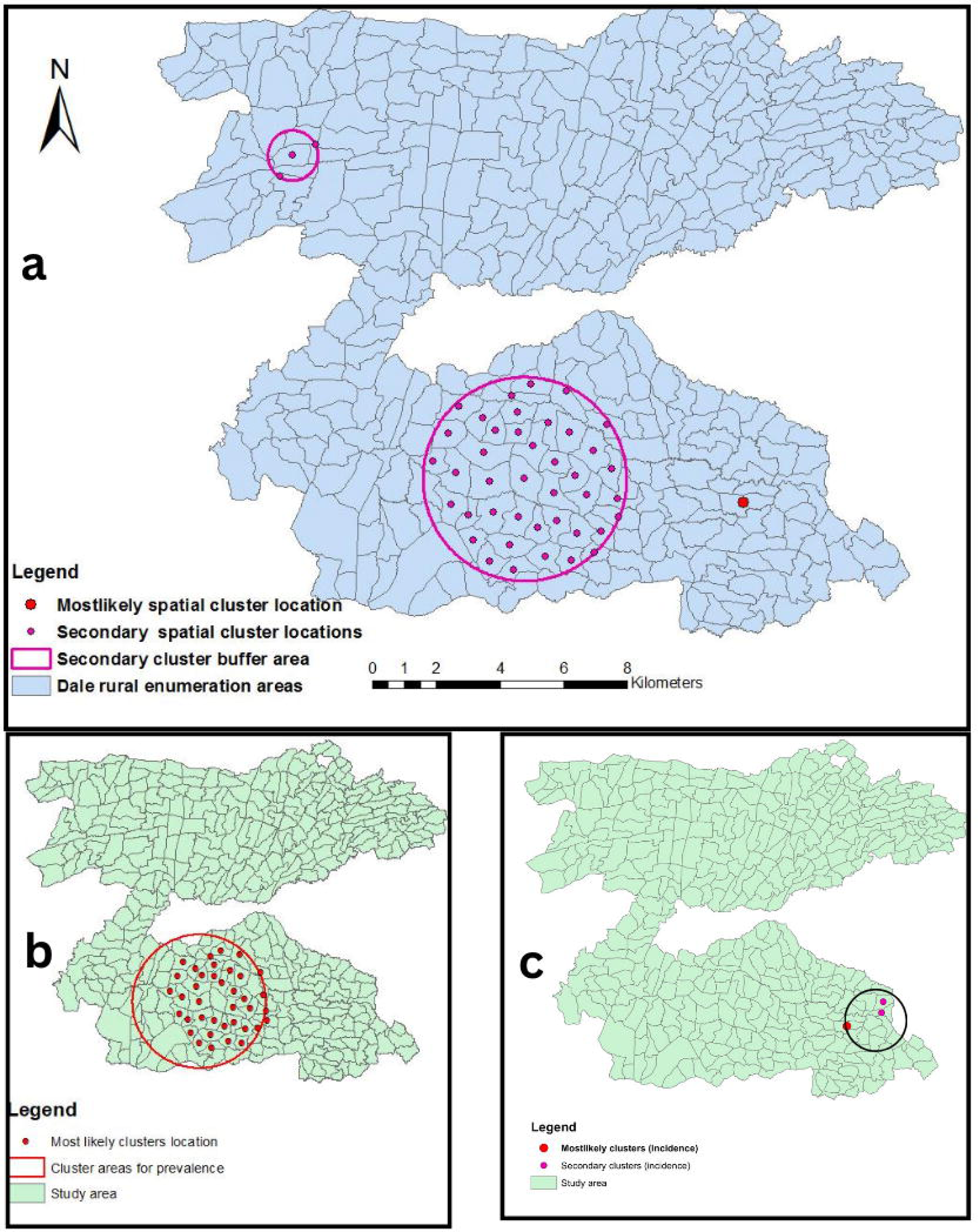
(a) Pulmonary tuberculosis clustering in the three rounds of household visits from October 2016 to September 2017; (b) Pulmonary tuberculosis clustering in the first round of household visits (prevalent cases) from October 2016 to January 2017; (c) Pulmonary tuberculosis clustering in the second and third rounds (incident cases) from February 2017 to September 2017.

**Table 2.** Results of cluster analysis of patients with pulmonary tuberculosis in Dale, Ethiopia, by scan statistics, in Dale, Ethiopia 2018.

| Spatial Clusters | Location | Observed cases (n) | Expected cases (n) | RR | Likelihood ratio | P |
| --- | --- | --- | --- | --- | --- | --- |
| Most likely cluster | 1 | 17 | 1.48 | 11.9 | 26.3 | <0.001 |
| Secondary clusters 1 | 41 | 50 | 23.7 | 2.09 | 15.4 | <0.001 |
| Secondary clusters 2 | 3 | 14 | 3.7 | 3.92 | 8.6 | 0.044 |

n, number; RR, relative risk. P, P-value

### Risk factors for PTB clustering

Table 3 shows the results of a multilevel analysis of the patients and EAs. EAs in PTB clusters were more likely far from health facilities than EAs not in PTB clusters. In our analysis, the PTB cluster was not associated with other factors examined in our regression model.

**Table 3.** Results of multilevel analysis: association between pulmonary tuberculosis clusters and risk factors in Dale, Ethiopia 2018.

| Covariates | Effect estimate |  | SE | z | p |
| --- | --- | --- | --- | --- | --- |
|  | Coefficient | 95% CI |  |  |  |
| (Intercept) | -25.5 | (-38.2, -12.6) | 6.5 | -3.9 | <0.001 |
| Women | -2.8 | (-8.0, 2.5) | 2.7 | -1 | 0.303 |
| Age ≥35 years | -0.5 | (-5.76, 4.82) | 2.7 | -0.2 | 0.862 |
| Ever married | -1.1 | (-5.8, 3.6) | 2.4 | -0.5 | 0.649 |
| Schooling (No) | -4.9 | (-10.47, 0.76) | 2.9 | -1.7 | 0.09 |
| BMI kg/m2 | 1.6 | (-0.57, 3.7) | 1.1 | 1.4 | 0.152 |
| <16 | 1,0 |  |  |  |  |
| 16-17 | -1 | (-3.59, 1.6) | 1.3 | -0.8 | 0.453 |
| 17-18.5 | 1.2 | (-0.73, 3.09) | 1 | 1.2 | 0.225 |
| ≥18.5 | -0.2 | (-2.76, 2.34) | 1.3 | -0.2 | 0.871 |
| MUAC in cm (>22) | -5.1 | (-13.36, 3.14) | 4.2 | -1.2 | 0.225 |
| Wealth index | -0.6 | (-1.6, 0.45) | 0.5 | -1.1 | 0.275 |
| Poor | 1 |  |  |  |  |
| Middle | -0.7 | (-5.92, 4.54) | 2.7 | -0.3 | 0.795 |
| Rich | -8.4 | (-16.96, 0.19) | 4.4 | -1.9 | 0.055 |
| BCG Scar (no) | 7.2 | (-5.75, 20.1) | 6.6 | 1.1 | 0.276 |
| *Number population ≥644 | -3.8 | (-9.36, 1.74) | 2.8 | -1.4 | 0.178 |
| *Distance in km from health centre ≥3 | 16.2 | (4.9, 27.5) | 5.8 | 2.8 | 0.005 |
| *Population density per square km<br>≥655 | 0.7 | (-4.15, 5.63) | 2.5 | 0.3 | 0.767 |
| *Altitude in metres ≥1793 | -3.8 | (-9.69, 2.18) | 3 | -1.2 | 0.215 |
| **Number of observation days | 0.1 | (-0.03, 0.05) | 0 | 0.7 | 0.506 |

cm, centimetre; kg, kilogram; km, kilometre; m<sup>2</sup>, meter square; SE, standard error; p, p-value \*These variables used cut-off above the mean values.\*\*continuous variable.

## Discussion

Based on active case-finding data, this study revealed clusters of symptomatic PTB cases in the Dale district in Ethiopia. In the southeast, northwest, and south-central parts of the district there was more PTB clustering, and the EAs with PTB clusters were farther from health centres. These areas may play an important role in transmission and suggest the need for targeted screening for early detection and treatment in areas with limited TB care. The study highlights the benefits of more active TB case-findings in these areas as majority of patients had not sought health services despite having symptoms (13, 27). Repeated active case-finding made TB care more accessible to everyone in the district. The strategy involves several rounds of screening, which may identify missing and subclinical TB cases in rural locations and improve our ability to pinpoint clusters since this technique basically removed most clusters in subsequent visits.

In program settings, the locations of cases are often linked to the diagnostic centres and not the patients’ households. Since many patients must travel to health centres for diagnosis, the notification data can, therefore, not always identify “real geographic” clusters, and units with “clusters” may include many visitors from other areas. Hence, the study findings from previous clustering studies using notification data, often reported in urban or semi-urban areas, where access to diagnosing and treating TB is less challenging (28,29).

Contrary to passive case-finding, active screening can be more costly but indicates clusters more likely to occur in areas farther from health centres. Such screening can help decentralize TB services, which may offer an effective means of improving access to early diagnosis, treatment and preventive therapy that helps long-term efforts to reduce TB clusters and infectious sources in the community (30, 31). Our case identification strategy did not depend on patient care seeking behaviour. Unlike previous studies that relied solely on passive case finding without accounting for undetected cases, we used trained HEWs who conducted regular screening visits on various health issues in their target population. These HEWs actively involve the entire population in the process, benefitting from their trust and familiarity with community screening. Expanding on this infrastructure, our study incorporated PTB symptom screening in all households within the district. The proportion of undetected symptomatic PTB cases is expected to be lower compared to the notification data from passive case-finding. While there may be a latency period between TB infection and symptom onset, repeated screening of the entire population over an extended period could help identify new cases previously asymptomatic who later become symptomatic as a disease progresses (13, 16). We used TB unit registrations to validate PTB cases and ensured that the identified patients received care and treatment.

In the multilevel analysis, we found that EAs in cluster areas were more distant from health facilities than other EAs; low TB notification and distance from the health facility were positively correlated (8). Distance from TB services affects access to TB services since it is interrelated with cost and time (32). Still, in rural settings, symptomatic people receive care late and with less sensitive diagnostic (13). TB prevention and care efforts should address patient delays and stigma affecting healthcare-seeking (33). TB screening targeted remote areas could detect clusters more reliably and reduce community transmission. There has been no previous research linking socioeconomic position and population clustering. In our analysis at the EA level, we sought not only to identify clustering but also risk factors for clustering that may require attention from public health practices. However, our findings did not show associations between clustering and wealth index and a multisite study would better identify socioeconomic-based differences in PTB clustering.

Our study has some limitations. We focused on patients with TB symptoms, missing the asymptomatic. While this may introduce bias for most individuals, this is usually a temporary condition, as symptoms in most will develop later (31, 34). Moreover, our study observed the populations for a year, with repeated screening of symptoms, so the number of undetected cases and asymptomatic individuals should be low. It’s important to note that misdiagnosis can occur despite the implementation of quality assurance measures, primarily due to the low sensitivity of smear microscopy. We did not include 17 PTB with missing location information in the study, which may influence identifying true clusters.

Populated urban areas have both more TB services and more TB contacts and transmission than rural areas, even though infection is often within close relatives in rural areas. Therefore, clusters of PTB cases may be more prevalent in urban areas where access to care is greater while being less reported in rural areas due to limited access to care (22). On the contrary, TB incidence has shown an inverse association with urbanization in developed, accessible health services and improved living standards. Identifying TB patients and clusters based only on passive case-finding misses undiagnosed and un-notified patients. GIS studies based on notifications identifying clusters cannot distinguish between “real” TB incidence differences and the role of health services (35).

In program context, many areas may perform well, but some areas with many cases and less developed systems may need more attention and focused strategies. GIS has the potential to be helpful in close contact and neighbourhood TB screening in areas where TB cases are concentrated (30) and indicate areas with low notification but with many cases as well. Active TB screening, on the other hand, can enhance TB programme outcomes in these areas but is costly and usually regarded almost impossible as a routine TB prevention and care strategy in limited-resource settings, and hence requires a thorough evaluation before implementation, particularly with X-ray and molecular diagnostic tests, which are very expensive. Therefore, a cheap regular HEW-based symptom screening, with routine monitoring of presumptive TB patients identified and confirmed TB among referred could help identify catchment facilities with lower than expected presumptive TB rates as low cluster areas may be more alarming than high cluster areas (36).

## Conclusions

The distribution of PTB was not uniform but concentrated in particular regions distant from health centres, indicating the need to strengthen decentralized TB services in remote settings. Routine systematic community screening using existing health infrastructure with HEWs may be costly but through targeted screening they can identify and refer persons with TB symptoms more quickly for diagnosis and treatment, thereby decreasing the transmission and contributing to the reduction of TB burden.

## Abbreviations

BCG, Bacillus-Calmette Guerin; BMI, Body mass index; EA, Enumeration area; GPS, Global positioning systems; GIS, Geographic information systems; HEW, Health extension worker; MUAC, Mid-upper arm circumference; PTB, pulmonary tuberculosis; TB, tuberculosis

## Ethics approval and consent to participate

Ethics approval was obtained from the AHRI-Alert Ethics Review Committee, Ethiopia (PO12/15), the National Research Ethics Committee, Ethiopia (no 104/2016), and the Regional Committees for Medical and Health Research Ethics in Norway (2015/1006). Written consent was obtained from participants and parent(s)/Guardian(s) for participants aged 15-17 years.

## Data sharing statement

Data cannot be shared publicly because of confidentiality to protect the study participantss since data sets generated and/or analysed in this study are not publicly available. Data are available upon reasonable request from the corresponding author, ABB. To receive access to the data, the applicant will need to provide ethical approval from IRB or an equivalent body and approval from the AHRI-ALERT and National Research Ethics Committees in Ethiopia.

## Author contributions

Conceptualization: ABB, BAW, DGD, EH; Data analysis: ABB, MHD; Methodology: ABB, BAW, MHD, SGH; Visualization: ABB, MHD; Writing original draft: ABB, MHD, BAW, SGH; Writing review and editing: ABB, MHD, DGD, EH, BAW, SGH. All authors reviewed and approved final manauscript.

## Data Availability

All data produced in the present study are available upon reasonable request to the authors

## Acknowledgements

We want to acknowledge, data collectors, healthcare workers and patients. We would like to thank the Sidama Regional Health Bureau for their support during the project period. The University of Bergen funded the publication fee.

## ORCID

Abiot Bezabeh Banti: https://orcid.org/0000-0003-0441-8845

Brita Askeland Winje: https://orcid.org/0000-0003-2858-7248

Sven Gudmund Hinderaker: https://orcid.org/0000-0003-2725-0248

Einar Heldal: https://orcid.org/0000-0002-8741-792X

## Supplemental appendix

Supplemantal file 1.

Supplemenatal file 2.

